# Can we Predict Training Performance with Shooting Heart Rate in Archers? – A Machine Learning Approach

**DOI:** 10.1101/2024.06.09.24308651

**Authors:** Chandra Sekara Guru, Uma Mahajan, Anup Krishnan, Karuna Datta, Deep Sharma

## Abstract

**Purpose:** Heart rate (HR) values during different phases of shooting can be used for performance analysis. Machine learning (ML) methods are used in predicting performance. We aimed to develop ML model to predict performance scores using shooting HR values and also to predict the importance of these parameters in an archer.

**Methods:** 32 archers (15 elite & 17 non-elite) shot two sessions of 30 arrows each indoor wearing heartrate chest monitor and were videographed. When each arrow was shot, 11 HR values were identified at different shooting phases. Other parameters with 35 linear variables and second-degree polynomial HR values were used to build ML models in Python V3.11.4. Session 1 and 2 total scores were used to train and test respectively. Root Mean Squared Error (RMSE) was used to evaluate the model performance after fine-tuning.

**Results:** RMSE of all 12 ML models ranged from 6.262 – 9.612. The Cat Boost model with the lowest RMSE of 6.262 was used to predict the Session 2 score. SHapley Additive exPlanations (SHAP) values showed each variable’s importance in prediction.

**Conclusions:** Sports age, resting systolic blood pressure, previous competition score, right hand grip-strength, age, HR before 2sec of arrow release, waist-to-hip ratio, concentration disruption trait anxiety and HR after 5sec of release are top parameters to predict score.

**Practical Applications:** ML model with shooting HR provides a better prediction of archery score of an individual archer.

## Introduction

Sports performance in archery is the ability of the archer to repeatedly shoot maximum score during the event (12). The complex interaction of the physiological, psychological and physical factors results in this coordinated motor performance in archery. Adaptations in these systems resulting from periodised scientific sports training can optimise the shooting performance in archery. Evaluation of the athletes periodically during the training phases is key to provide insight to the coach and performance staff about the effect of the training imparted. Analysis of these performance parameters is used by the coaching personnel to undertake corrective action in training for optimum performance.

Various biophysical parameters have been used to predict performance in archery. One such performance parameter is the shooting heart rate which is considered as an outcome effect of the adaptive responses of the physiological and the psychological systems. Most studies have captured the heart rate values before shooting and correlated with the archery scores (1,3). Given the importance of the key phases of shooting i.e. aiming, release and follow through and correlation of physiological-biomechanical parameters recorded during these phases with archery score, we intended to capture the heart rate values during these phases for deeper insights (10,15,23). Even though heart rate values during shooting have been correlated with the performance score, shooting heart rate values alone without the other performance factors may not aid in performance prediction and training optimisation (1,8,12,20).

Moreover, every athlete is unique given the differences in genetic composition, gender, anthropometric characteristics and psychological trait. Specific determinants and motor skills required as per the demand of the sport further add to the complexity. In addition, the parameters that are periodically evaluated are closely inter-related and stochastic in nature. Hence research studies incorporating Machine

Learning (ML) regression models have been postulated instead of routine statistical regression methods to classify the archers based on their performance prediction (14,16,17,25).

Anthropometric characteristics, fitness parameters, physiological resting variables and biomechanical variables are used to develop the ML prediction models to either identify or classify the potential archers (14,16,17,24,25). There is lack of a performance predictive ML model incorporating both the key performance parameters and the heart rate values captured during shooting. Moreover, understanding the individual parameter’s predictive importance is the key for optimising the performance in a specific archer. Such a ML model can aid the coach to manoeuvre the training methods to provide individualised performance enhancement.

The aim of this study was to develop a machine learning model to predict the archery performance score with key performance parameters including the heart rate values during key phases of shooting and determine the predictor importance of such performance parameters for the individual archer in order to provide individualised training.

### Materials & Methods

An observational study was designed among the male archers of a residential sports institute in western Maharashtra, India after obtaining necessary ethical clearance from the Institutional ethical committee (IEC/NOV/2016). 32 Indian archers, i.e. 15 elite and 17 non-elite, volunteered to take part in this study after providing informed consent. There were 21 recurve and 11 compound archers in total. All participants were into active sports training and participated in the absence of any audiences or coaching personnel.

The study involved shooting two sessions of 30 arrows each comprising of 10 ends over each session from a distance of 18m in an indoor archery hall as per the World Archery Association standard rules (8). The entire process of shooting and corresponding scores by each arrow were recorded by the principal investigator. The environment set-up and pre-participation requirements were same for all the participants. Heart rate was measured using a heart rate monitor with chest belt (Polar Electro Oy, Kemple, Finland). The wrist watch displaying the heart rate was fixed to a tripod and filmed simultaneously during the shooting during the entire study. The video recorded was set at 50 frames per second (HDR-PJ670, Sony Electronics, San Diego, CA, USA).

Slow motion analysis of the recordings were done to identify the time of release of the arrows. A total of 10 frames, each 5 seconds before and after the arrow release of each arrow were also identified by the principal investigator. All the frames had the shooting heart rate value displayed on the watch. Thus, a total of 11 shooting heart rate values covering the aiming, release and follow through phase were captured for each arrows. The detailed materials and methods has already been presented in the pilot study by our team already (8). With a total of 32 archers shooting 60 arrows each and 11 shooting heart rate values, the total heart rate values sample used in this study to develop and test the machine learning performance predictive model was 21,120. The mean heart rate values were plotted against the 11 frames depicting the three archery phases i.e. aiming (-5 sec to -1 sec), release (0 sec), follow through (+1 sec to +5 sec) to identify the pattern.

Descriptive analytics namely frequencies (n), percentages (%) and mean, standard deviation along with 95% confidence intervals (95%CI) were used to summarize categorical and numerical data respectively. Heat map was used to assess the correlation between numeric variables. The category of archers grouped based on the type of archery and level of the archers. i.e. elite vs non-elite and compound vs recurve, relevant biophysiological variables like anthropometric characteristics, physiological parameters, psychological variables, subjective sleep quality rating, shooting heart rate values and sports training parameters like sports age, phase of training, previous competition performance were used as the independent predictor variables (See Supplementary table 1). The archery score out of 300 in session1 and session2 were used as the outcome variables to build the ML regression models.

A total of 12 ML regression models were developed using two approaches due to the curvilinear nature of the shooting heart rate values observed when plotted against time (Figure 1). We assumed all the 35 parameters as linear variables in the first approach to develop 06 ML models. Whereas in the second approach, in addition to assuming all as linear variables, we also considered the second degree polynomials of shooting heart rate values (11 heart rate values), thus making a total of 46 variables in each of the ML models. Session1 data was used to train the ML models. These ML Models were fine-tuned using ‘Grid Search CV’ cross-validation. The predictive performance of each ML model was tested using the Session2 score dataset.

**Figure 1:**
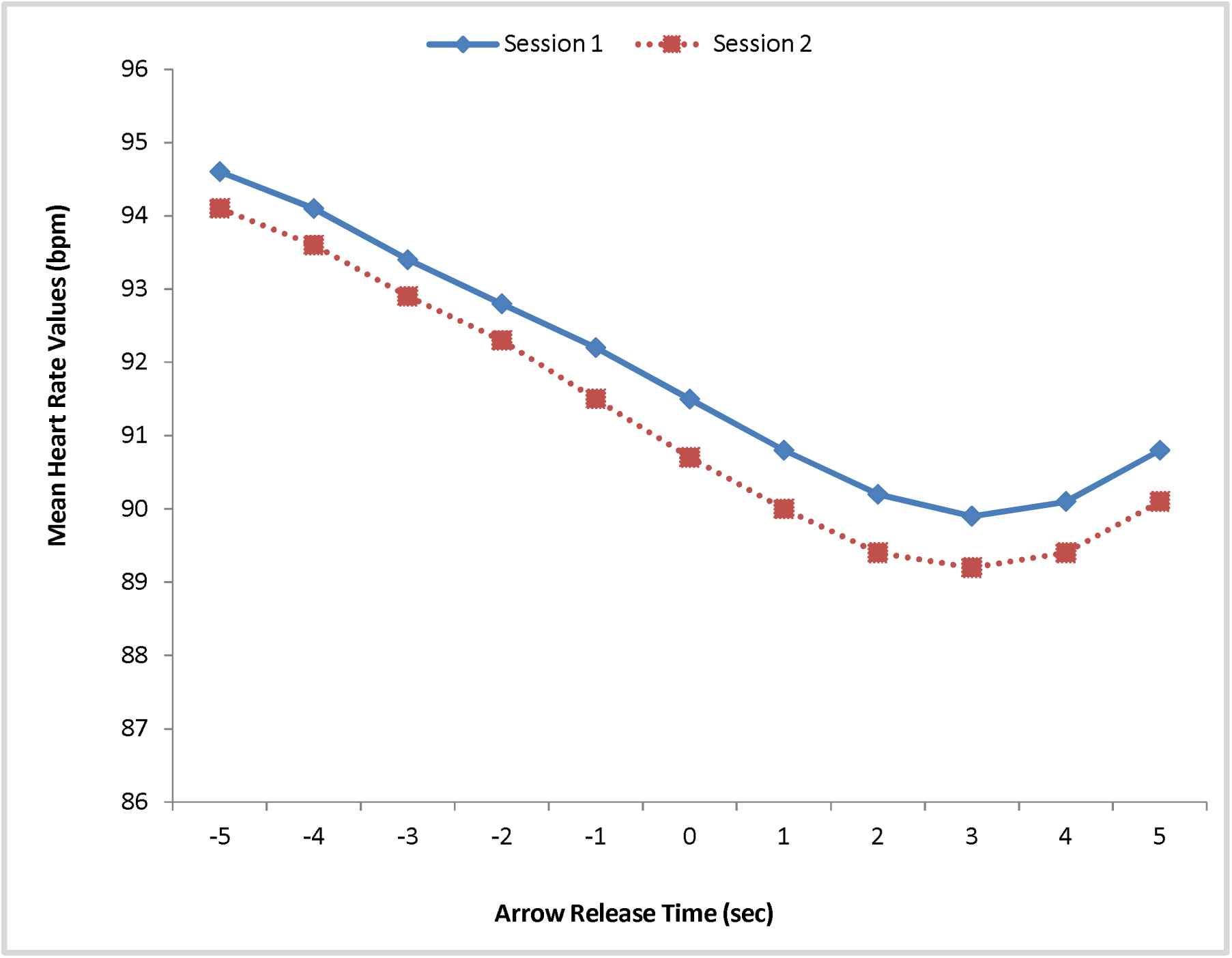
**Heart Rate Values in Indian Archers**

Root Mean Squared Error (RMSE) was derived for evaluating the model performance. The ML model with the lowest RMSE was considered as the final model. ‘SHapley Additive exPlanations (SHAP)’ values were calculated for the final ML model to evaluate the variable importance of each variable in predicting the archery score. All the analysis and ML modelling were done in Python v3.11.4

## Results

A total of 32 male archers with mean age of 21.8 years and mean sports experience of 7.6 years participated in this study. The demographic and sports profile of the participants are tabulated in Table 1. The mean shooting heart rate values identified during the aiming, release and follow through phases showed a specific deceleration pattern in both the sessions (Figure 1). Maximum deceleration of the mean shooting heart rate values was observed during +2 and +3 seconds after release of the arrows in the follow through phase and then gradually started to increase further. Even though the mean heart rate values were lower in session 2 compared to the session 1, the mean archery scores in both sessions did not differ.

**Table 1:** Profile of Indian Archers.

| Variables | Total | Variables | Total |
| --- | --- | --- | --- |
|  | (N=32) |  | (N=32) |
|  |  | Anthropometric & Sports related |  |
| Archer Group |  |  |  |
| Elite compound | 7 (21.9%) | Age (years) | 21.8 (19.77, 23.83) |
| Elite recurve | 8 (25.0%) | Sports age (years) | 7.6 (5.58, 9.52) |
| Non-elite compound | 4 (12.5%) | Training age (years) | 4.1 (2.79, 5.41) |
| Non-elite recurve | 13 (40.6%) | Height (cm) | 168.7 (166.37, 170.94) |
| Session 1: Arrow release time (sec) | Heart rate values (bpm) | Weight (kg) | 66.7 (63.12, 70.19) |
| -5 | 94.6 (90.29, 98.92) | Waist/hip ratio | 0.9 (0.85, 0.88) |
| -4 | 94.1 (89.72, 98.43) | Body fat (%) | 10.9 (9.84, 11.92) |
| -3 | 93.4 (89.03, 97.85) | Physiological |  |
| -2 | 92.8 (88.37, 97.25) | Right hand grip strength | 41.6 (39.01, 44.11) |
| -1 | 92.2 (87.70, 96.64) | Left hand grip strength | 40.8 (38.20, 43.40) |
| 0 | 91.5 (87.03, 96.00) | Resting heart rate (bpm) | 58.8 (56.10, 61.59) |
| 1 | 90.8 (86.30, 95.30) | Resting blood pressure - Systolic (mmHg) | 114.5 (111.16, 117.77) |
| 2 | 90.2 (85.65, 94.67) | Resting blood pressure -<br>Diastolic (mmHg) | 64.4 (61.73, 67.02) |
| 3 | 89.9 (85.40, 94.47) | Heart rate before Warm-up | 88.9 (85.07, 92.74) |
| 4 | 90.1 (85.49, 94.67) | **Psychological** |  |
| 5 | 90.8 (86.15, 95.50) | Somatic trait anxiety (9-36) | 13.9 (12.97, 14.90) |
| **Session 2: Arrow** |  |  |  |
| **release time (sec)** |  | Cognitive Trait Anxiety (7-28) | 13.1 (11.64, 14.48) |
|  |  | Concentration Disruption Trait<br>anxiety (5-20) | 8.2 (7.45, 8.98) |
| -5 | 94.1 (90.12, 98.15) |  |  |
| -4 | 93.6 (89.54, 97.65) | **Sleep quality [n (%)]** |  |
| -3 | 92.9 (88.85, 97.05) | Good | 25 (78.1%) |
| -2 | 92.3 (88.12, 96.41) | Average | 7 (21.9%) |
| -1 | 91.5 (87.32, 95.72) | **Highest performance in previous competition** |  |
| 0 | 90.7 (86.49, 95.00) | Archery score (out of 720) | 660.4 (649.74, 671.14) |
| 1 | 90.0 (85.75, 94.34) | **Type of previous highest performing competition** |  |
| 2 | 89.4 (85.10, 93.72) | International | 7 (21.9%) |
| 3 | 89.2 (84.83, 93.47) | National | 11 (34.4%) |
| 4 | 89.4 (84.98, 93.75) | Below national | 14 (43.8%) |
| 5 | 90.1 (85.59, 94.55) |  |  |
| **Training phase** |  |  |  |
| **[n (%)]** |  | **Performance Outcome** | **Archery scores** |
| Preparatory | 24 (75.0%) | Session 1 score (out of 300) | 279.4 (276.70, 282.05) |
| Pre-competition | 8 (25.0%) | Session 1 score (out of 300) | 279.5 (276.01, 283.05) |

Note: Values are expressed as Mean (95% Confidence Interval)

Correlation heatmap of predictor variables and the archery score is shown in Figure 2. The blue and red colour boxes represent the positive and negative correlation respectively. The lighter to darker shade represent the strength of the correlation from low to high. Highest archery score in previous competition, age of the archer, training age at the institute, sports age (experience) were positively correlated with the session 1 archery score. The shooting heart rate values identified during all three phases i.e. aiming, release and follow through were negatively correlated with the archery score.

**Figure 2:**
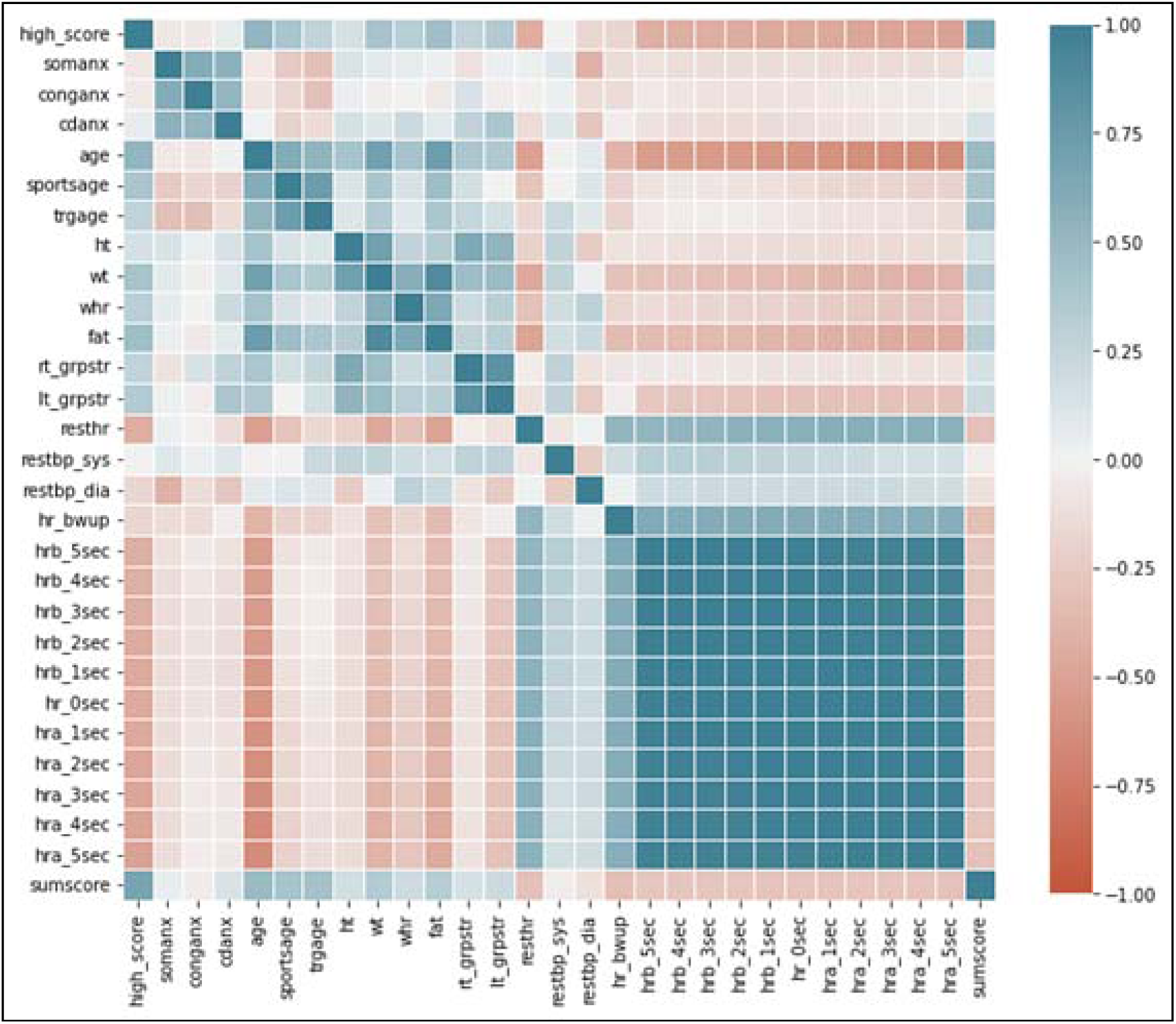
**Correlation Heatmap of Predictors Variables and Archery Scores**

ML models namely linear regression, polynomial regression, elastic net, random forest, extreme gradient (XG) boosting, categorical (Cat) boosting, light gradient boosting machine (GBM) were developed using both the approaches i.e. linear and polynomial features. Table 2 shows the ML models developed using both the approaches and their respective RMSE along with the best model parameters. The linear Categorical (Cat) Boost ML model was chosen as the final model with the lowest RMSE of 6.262 among the 12 developed models after testing with session 2 dataset. The Cat boost linear ML model developed using session 1 dataset was used to predict the archery score and was tested with the actual session 2 archery score of all the archers individually (Figure 3). The average predicted score using Cat Boost linear ML model was 279.4. Figure 3 depicts the difference in the actual and predicted archery scores as well as the deviation away from the average predicted score between the category groups of the archers especially between elite and non-elite archers.

**Figure 3:**
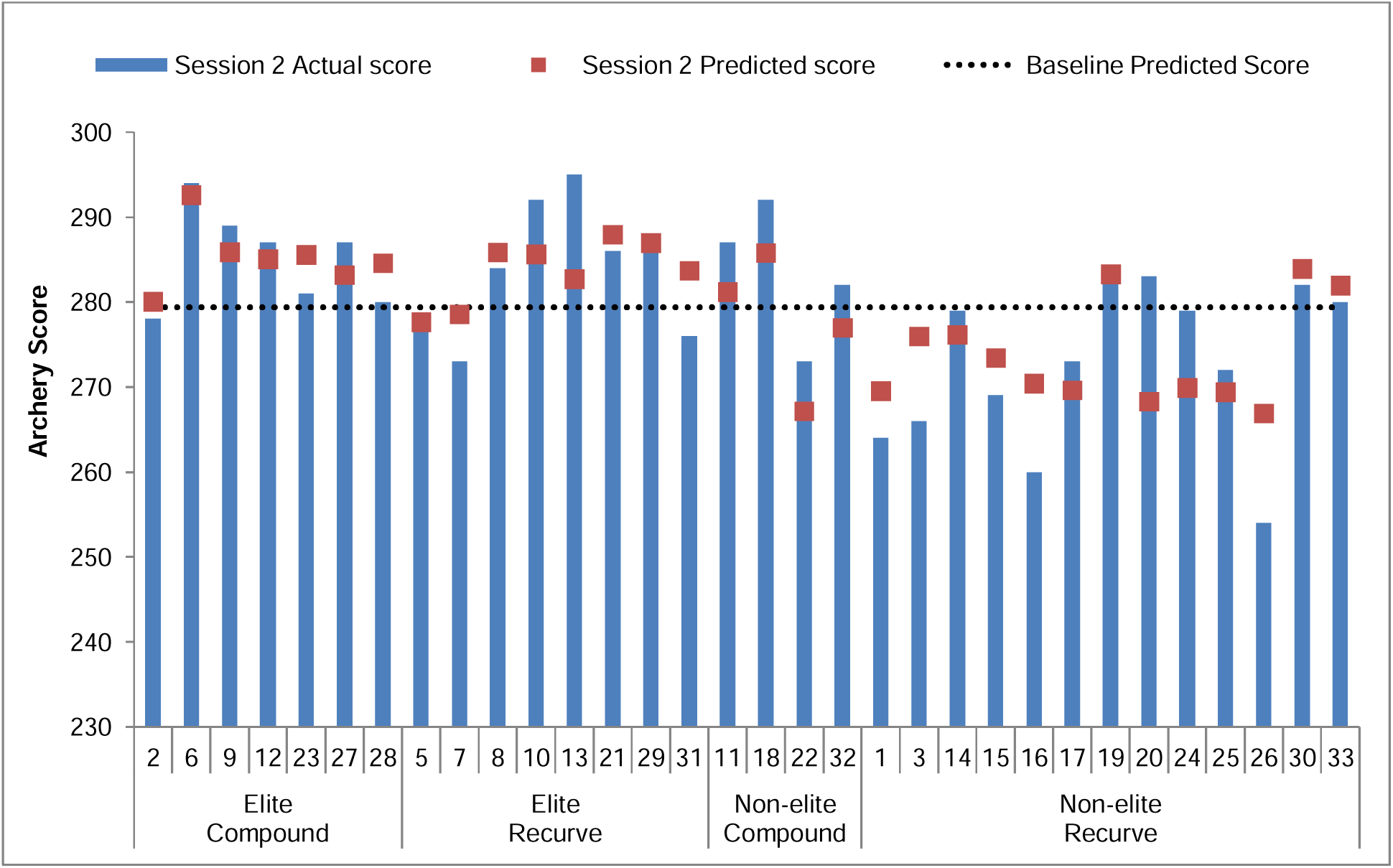
**Session 2 Actual and predicted archery scores (CatBoost model – 35 variables)**

**Table 2:** Machine Learning Model Performance in Testing Dataset.

| Models | RMSE | Best parameters |
| --- | --- | --- |
| **With linear features (35 variables)** |  |  |
| Linear regression | 7.489 |  |
| Elastic Net | 7.606 | alpha: 25, l1_ratio: 0.5 |
| Random forest | 6.919 | max_features: 5 |
| XG Boost | 6.728 | learning_rate: 0.15, max_depth: 2, n_estimators: 40 |
| Cat Boost | 6.262 | learning_rate: 0.5, max_depth: 4, n_estimators: 25 |
| Light GBM | 9.612 | learning_rate: 0.1, max_depth: 2, n_estimators: 20 |
| **With polynomial features (46 variables)** |  |  |
| Polynomial regression | 8.325 |  |
| Elastic Net | 7.394 | alpha: 15, l1_ratio: 0.5 |
| Random forest | 6.852 | max_features: 6 |
| XG Boost | 6.728 | learning_rate: 0.15, max_depth: 2, n_estimators: 40 |
| Cat Boost | 7.334 | learning_rate: 0.15, max_depth: 2, n_estimators: 20 |
| Light GBM | 9.612 | learning_rate: 0.1, max_depth: 2, n_estimators: 20 |

Note: RMSE = Root mean squared error

### Predictor importance of the Bio-Physiological variables

Figure 4 shows the summary plot of the SHAP values derived using the Cat boost ML model with linear shooting heart rate values. The SHAP values depict the overall importance of the independent variables in predicting the archery performance score in the session 2 dataset (Figure 4). The predictor/independent variable names are shown on Y-axis and SHAP values of each variable are shown on X-axis in the plot. The variables are displayed from top to bottom in order of their importance from most important to least important in predicting the archery score respectively. The SHAP values from low to high are shown using blue and red colour respectively. The lighter to darker shade represents the magnitude of the SHAP values from low to high. Each point on each predictor variable represents the SHAP value of each archer. Figure 5 shows the difference in the predictor variables contribution using SHAP analysis of the Cat Boost ML model between archers from the same categories i.e. elite compound (archer ID 6 vs archer ID 23) and non-elite recurve (archer ID 20 vs 19).

**Figure 4:**
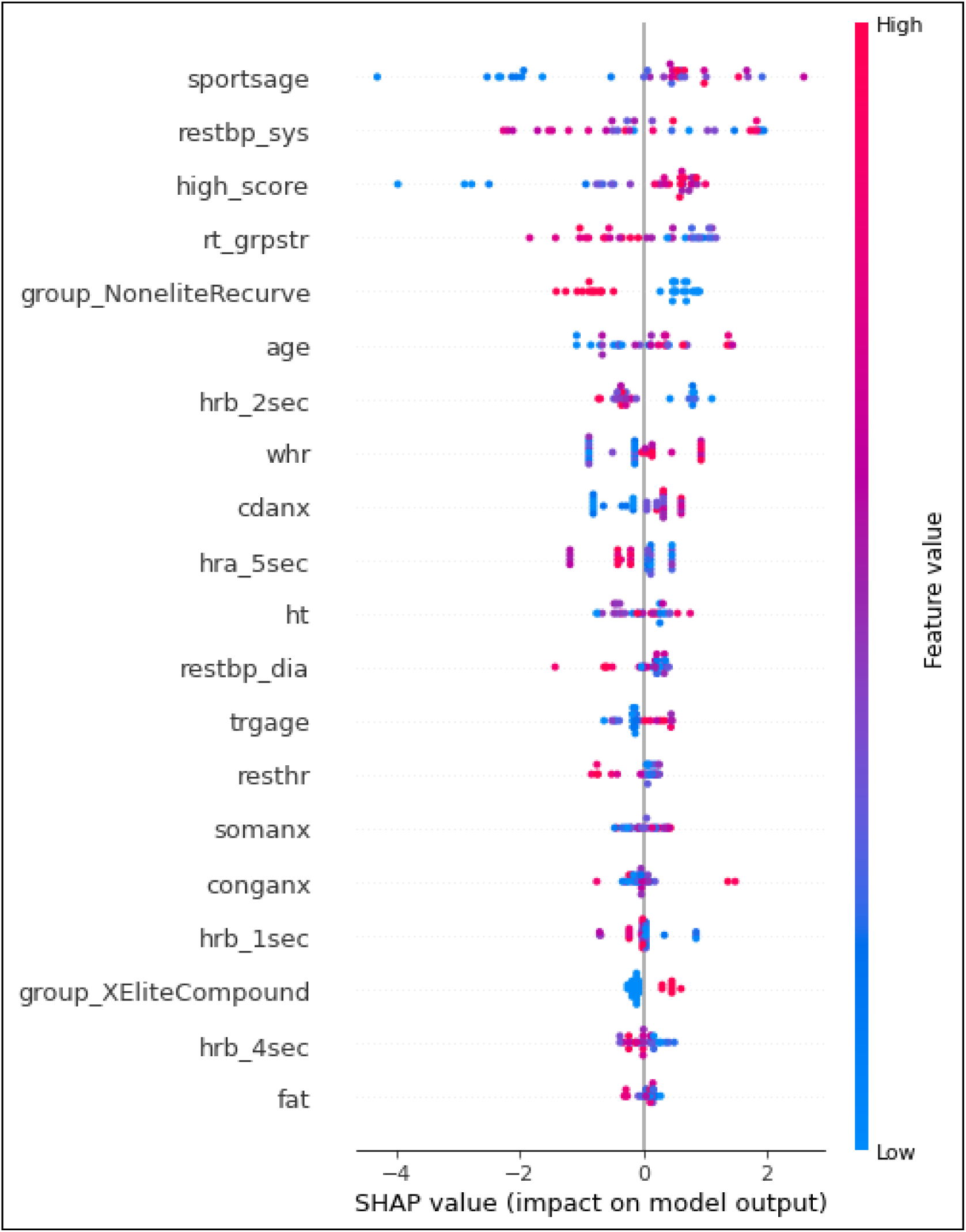
**Predictor Importance of Cat Boost Model using SHAP Analysis**

**Figure 5:**
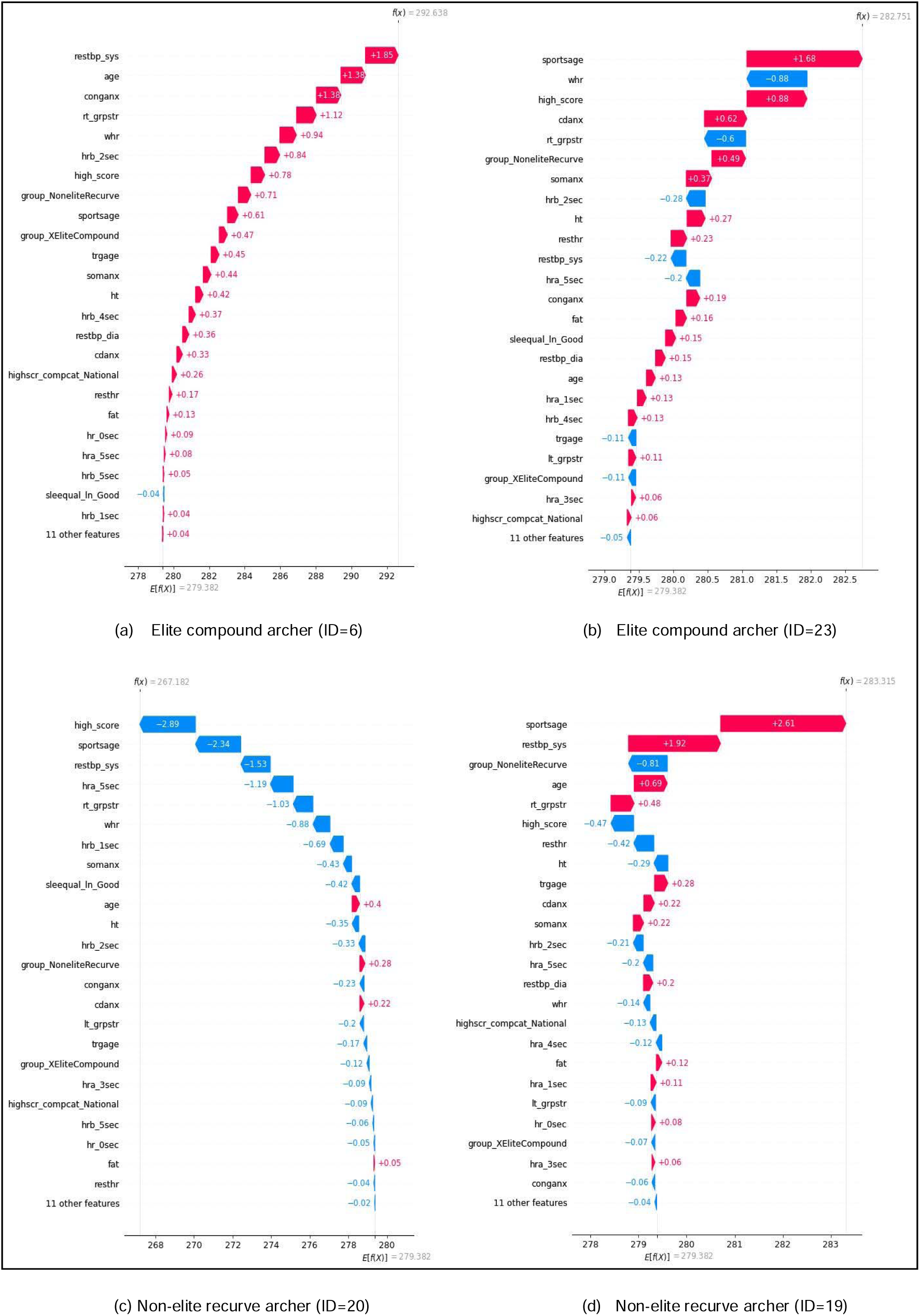
**Individual Archer-level Importance of Predictors**

## Discussion

We aimed at measuring the heart rate during the key phases of shooting using basic field-side aids to highlight that this method can be easily incorporated by coaches and scientists during routine periodic evaluation (8). Our attempt to use multiple variables along with shooting heart rate to develop a performance prediction ML model for personalised training of the athlete is a novel approach.

The performance variables correlated with archery scores are varied, generally grouped under physical, mechanical, physiological and psychological components. These factors vary for different events and their component contribution also may vary between individual athletes (11). The individual archer may develop unique strategies and adaptations to training in these performance components (26). Hence we included four categories of archers in our study to gain deeper insight pertaining to the difference in these performance parameters. The grouping was done based on competitive level of archers who participated in international competition or National camp and below (elite vs non-elite), and archery event (recurve vs compound). A total of 35 variables under anthropometric, sports related, physiological, psychological, sleep and shooting heart rate values during the key phases of shooting were included based on previous research studies and correlation with archery performance scores (Supplementary Table 1) (1,3,4,7,8,11,14).

### Pattern of Shooting Heart rate values

A unique feature selection variable to predict performance included in this study was shooting heart rate values during the three main shooting phases. Clemente et al and Aggarwala et al have found an inverse correlation between the pre-shooting heart rate values and the archery scores in their studies (1,3). Robazza et al in his study has identified a specific deceleration pattern of heart rate during the transition from draw-aiming to release phase which correlated with the archery scores and level of archers (19). In our study, we found that the shooting heart rate values showed a specific deceleration pattern which extended beyond the release phase into the follow through phase i.e. +2 and +3 seconds (Figure 1).

The deceleration pattern in shooting heart rate values has been attributed to the outcome effect of coordinated motor, breathing, attention and visual responses during the aiming - shooting phases and postulated due to the cardiac-somatic coupling, intake-rejection hypothesis or the quiet eye phenomenon in aiming sports like golf, archery and shooting (6,18,19). The specific pattern of heart rate deceleration can thus provide better clues to coaches for planning sports training. The deceleration pattern when plotted against the time (11 frames of the shooting phases) depicted a curvilinear graph (Figure 1). Hence, we included the second degree polynomials of this mean shooting heart rate values in our second approach to develop the ML model, so that this aspect may improve the accuracy of prediction.

### Machine learning model for performance prediction

The performance components are multiple and are multicollinear in nature, especially the physical, physiological and psychological variables given the inter dependency between the organ systems during execution of a motor task (14). This can be clearly highlighted in the correlation heatmap plot of the predictor variables with archery score (Figure 2). Moreover, methods using traditional statistical regression filter these complex inter dependent variables during feature selection analysis and hence may have limitations in performance prediction. Hence in recent studies, machine learning models have been evaluated to tackle the more complex performance variables that predict sports performance and identify potential athletes (5,9,13,17,25).

In our study, we used 12 different ML models using two different approaches and the RMSE ranged between 6.262 and 9.612 for all models Even though we used second degree polynomials, the Cat Boost ML model with linear heart rate values itself predicted the archery score better when tested showing the lowest RMSE of 6.262 (Table 2).. The Cat Boost model (linear 35 variables) with lowest RMSE of 6.262 was used to predict the Session 2 score and tested against the actual session 2 scores (Figure 3), which is again an important strength of this study given the large data sets for both training and testing.

The predicted scores for the individual athletes using the Cat Boost ML model are compared with actual scores made in session 2 (Figure 3). The Cat Boost linear ML model predicted a baseline archery score of 279.38 out of 300 in session 2 across all groups of archers. An interesting aspect is the difference in deviation of predicted as well as actual scores from the baseline predicted score noted between the archer groups. Among most of the non-elite recurve archers, the actual as well as predicted scores were below the baseline predicted score which might been due to the difference in the learning curve and comparatively tougher equipment handling technique involved in recurve archery (2). This gives an insight to the coach on the training aspect required for the individual archer to cover up for the baseline predicted score.

### Importance of the performance parameter in predicting the score

The input of baseline predicted score alone may give a general idea to the coach and the athlete for further performance enhancement. The relative contribution of various individual variables to the predicted score can aid in personalised training. Hence, SHAP analysis on the Cat Boost model was done to identify the contribution of each independent variable in predicting the performance (Figure 4). Sports age, resting systolic blood pressure, highest archery score by the archer in previous competitions, right hand grip strength, non-elite recurve group, age, heart rate value +2 sec of arrow release, waist-to-hip ratio, concentration disruption trait anxiety, and heart rate value +5 sec of arrow release were the top 10 important variables which played vital role in prediction of the archery score.

Similar to our findings, Muza and Taha et al have also highlighted that the lower resting blood pressure, resting heart rate, and higher handgrip strength as performance indicators of high performing potential archers (17,25). Similarly, Uma et al also have found that maximum handgrip strength, higher fat mass, and age are important features in their machine learning feature selection analysis for archery performance (14).

Our study, in addition to these variables has identified concentration disruption trait anxiety, a component of the Sports anxiety scale in addition to the cognitive trait anxiety and somatic anxiety trait. The concentration disruption trait score ranges between 5 and 20 and a higher score is negatively correlated with performance (22). In our study the mean score was only 8.2 with 95% confidence interval of 7.45 to 8.98 (Table 1). Shaari et al, in their study among 44 National level archers have also found a similar concentration disruption mean score of 10.33 (21).

Further, as discussed earlier, our study has included the shooting heart rate during key phases. The SHAP analysis further amplified the importance of the shooting heart rate values during follow-through phase at +2 and +5 seconds after the release of the arrows in predicting the archery score. This is an important input for coaches to focus during the training sessions. Also, sports related variables like sports age (experience), previous high score in competitions and non-elite recurve archers are key factors that predict performance.

### Predictor importance for planning personalised training

The contribution of the predictor variables for each archer in predicting the archer performance varies. Information on contribution these variables in predicting the performance for that particular archer will provide better insight for the coaches to plan personalised training and enhance their performance. A comparison of how the variables contributed to the actual score from the baseline predicted score between two archers of same group namely Elite compound (Figure 5a and 5b) and Non elite recurve (Figure 5c and 5d) are depicted.

In case of the elite compound archer-ID 6, the resting systolic blood pressure contributed approximately 2 points (+1.85) from the average predicted score of 279 to improve up to 292 (Figure 5a). Whereas in case of the elite compound archer-ID 23, the sports age, in other words experience of the archer contributed approximately 2 points (+1.68) from the average predicted score of 279 to improve up to 282 (Figure 5b). Age, the cognitive trait anxiety, right hand grip strength, waist-to-hip ratio, and heart rate before 2sec of arrow release improved the predicted score by 1 point each in archer ID 6 (Figure 5a) whereas waist-to-hip ratio reduced the prediction by 1 point score in archer ID 23 (Figure 5b).

However, in non-elite recurve archer ID 20, the predicted score decreased to 267 from the average predicted score of 279. Previous competition performance, sports age, resting systolic blood pressure, heart rate value at -5 sec of arrow release, and right hand grip strength played major role in decreasing the predictions by approximately 9 points (Figure 5c). In non-elite recurve archer ID 19, the predictions further improved to 283 from the average predicted score of 279. Sports age and resting systolic blood pressure were vital in improving the predicted archery score by 3-4 points (Figure 5d). Thus within the same archer group, it can be seen that the contribution of the predictor features differs which may be a clue to the coach for planning personalised training.

## Conclusion

As the performance variables are varied and collinear, machine learning model pose a better option than traditional statistical prediction models. Out of the 12 tested ML models, the Cat Boost model has the lowest RMSE predicting a baseline archery score of 279 in session 2. SHAP analysis of the Cat Boost model provided the contribution of various variables in the predicting the performance which varies with individual athletes. Inclusion of physical fitness variables to the model in future may increase the accuracy of prediction. The ML model predicted has been trained and tested for indoor archery setting in a controlled environment. Similar studies in the future with competition setting as per the archery group in large sample may be incorporated for developing appropriate performance prediction models.

### Practical Application

- In addition to the resting physiological values, including shooting heart rate values during different phases highlights the complex motor task execution involved in archery and pave way for better training programming and monitoring.
- Cat Boost Linear machine learning model developed using the anthropometric-biophysiological variables has the least root mean square in predicting baseline archery scores in an indoor setting. The difference in predicted and actual score can aid the coaches to tailor the training program.
- SHAP values calculated using the Cat Boost ML provide the predictor importance of the performance variables which again can provide deeper insights for an individual archer and help performance team to provide personalised inputs to the coach and athletes.

## Data Availability

All data produced in the present study are available upon reasonable request to the authors

## Acknowledgements

The authors express their gratitude to the coaches and support staff of the Army Sports Institute, Pune, India, for their support during the conduct of the study.

