## Supplementary table 1 for "Can we Predict Training Performance with Shooting Heart Rate in Archers? – A Machine Learning Approach"

**Supplementary Table 1: Variable Names and Labels**

| **Variable name** | **Type of variable** | **Variable label** |
| --- | --- | --- |
| **Predictors/ Independent variables** | |  |
| group_NoneliteRecurve | Binary | Group - Non elite recurve (Ref: Non elite compound) |
| group_XEliteCompound | Binary | Group - Elite compound (Ref: Non elite compound) |
| group_XEliteRecurve | Binary | Group - Elite recurve (Ref: Non elite compound) |
| hrb_5sec | Numeric | Heart rate -5 sec arrow release |
| hrb_4sec | Numeric | Heart rate -4 sec arrow release |
| hrb_3sec | Numeric | Heart rate -3 sec arrow release |
| hrb_2sec | Numeric | Heart rate -2 sec arrow release |
| hrb_1sec | Numeric | Heart rate -1 sec arrow release |
| hr_0sec | Numeric | Heart rate 0 sec arrow release |
| hra_1sec | Numeric | Heart rate +1 sec arrow release |
| hra_2sec | Numeric | Heart rate +2 sec arrow release |
| hra_3sec | Numeric | Heart rate +3 sec arrow release |
| hra_4sec | Numeric | Heart rate +4 sec arrow release |
| hra_5sec | Numeric | Heart rate +5 sec arrow release |
| trgphase_Xprecompetition | Binary | Training phase - Pre competition |
| age | Numeric | Age (years) |
| sportsage | Numeric | Sports age (years) |
| trgage | Numeric | Training age (years) |
| ht | Numeric | Height (cm) |
| wt | Numeric | Weight (kg) |
| whr | Numeric | Waist-to-hip ratio |
| fat | Numeric | Body fat (%) |
| rt_grpstr | Numeric | Right hand grip strength |
| lt_grpstr | Numeric | Left hand grip strength |
| resthr | Numeric | Resting heart rate (bpm) |
| restbp_sys | Numeric | Resting blood pressure - Systolic (mmHg) |
| restbp_dia | Numeric | Resting blood pressure - Diastolic (mmHg) |
| hr_bwup | Numeric | Heart rate before Warm-up |
| somanx | Numeric | Somatic trait anxiety (9-36) |
| conganx | Numeric | Cognitive trait Anxiety (7-28) |
| cdanx | Numeric | Concentration disruption trait anxiety(5-20) |
| sleequal_ln_Good | Binary | Sleep quality last night - Good (Ref: Average) |
| high_score | Numeric | Highest score in previous competitions |
| highscr_compcat_International | Binary | Type of previous highest performing competitions - International (Ref: Below national) |
| highscr_compcat_National | Binary | Type of previous highest performing competitions - National (Ref: Below national) |
| **Dependent/ outcome variable** | |  |
| sumscore | Numeric | Total archery score in Session 1, Session 2 |
